# High effectiveness of multimodal infection control interventions in preventing SARS-CoV-2 infections in healthcare professionals: a prospective longitudinal seroconversion study

**DOI:** 10.1101/2020.07.31.20165936

**Authors:** Thomas Theo Brehm, Dorothee Schwinge, Sibylle Lampalzer, Veronika Schlicker, Julia Küchen, Michelle Thompson, Felix Ullrich, Samuel Huber, Stefan Schmiedel, Marylyn M Addo, Marc Lütgehetmann, Johannes K Knobloch, Julian Schulze zur Wiesch, Ansgar W Lohse

## Abstract

**Objective:** To assess the effectiveness of multimodal infection control interventions in the prevention of SARS-CoV-2 infections in healthcare professionals

**Design:** Sequential follow-up study

**Setting:** Largest tertiary care centre in northern Germany

**Participants:** 1253 employees of the University Medical Center Hamburg-Eppendorf were sequentially assessed for the presence of SARS-CoV-2 IgG antibodies at the beginning of the covid-19 epidemic (20 March – 9 April), one month (20 April – 8 May), and another two months later (22 June – 24 July). Of those, 1026 were healthcare workers (HCWs) of whom 292 were directly involved in the care of covid-19 patients. During the study period, infection control interventions were deployed, those included i) strict barrier nursing of all known covid-19 patients including FFP2 (N95) masks, goggles, gloves, hoods and protective gowns, ii) visitor restrictions with access control at all hospital entries, iii) mandatory wearing of disposable face masks in all clinical settings, and iv) universal RT-PCR admission screening of patients.

**Main Outcome Measures:** SARS-CoV-2 IgG seroconversion rate

**Results:** At the initial screening, ten participants displayed significant IgG antibody ratios. Another ten individuals showed seroconversion at the second time point one month later, only two further participants seroconverted during the subsequent two months. The overall SARS-CoV-2 seroprevalence in the study cohort at the last follow-up was 1.8%, the seroconversion rate dropped from 0.81% to 0.08% per month despite a longer observation period. Amongst HCWs seropositivity was increased in those directly involved in the care of patients with SARS-CoV-2 infections (3.8%, n=11) compared to other HCWs (1.4%, n=10, P=0.025). However, after the adoption of all multimodal infection control interventions seroconversions were observed in only two more HCWs, neither of whom were involved in inpatient care.

**Conclusion:** Multimodal infection control and prevention interventions are highly effective in mitigating SARS-CoV-2 infections of healthcare professionals.

## INTRODUCTION

Healthcare workers (HCWs) are at the front line of the coronavirus disease 2019 (covid-19) pandemic response and disproportionally at risk of contracting severe acute respiratory syndrome coronavirus 2 (SARS-CoV-2) due to occupational exposure.^1 2^ The Chinese ophthalmologist Li Wenliang, who was one of the first physicians to issue emergency warnings about a novel viral pneumonia that was later identified as covid-19 died after being infected when caring for a pre-symptomatic patient. ^3^ This individual case as well as larger case series illustrate the considerable challenges of HCW protection, especially since the risk of infections from pre- or asymptomatic SARS-CoV-2 infected individuals was initially often underestimated.^4 5^ Protection of HCWs from infection requires not only strict application of personal protective equipment (PPE), but also the prompt identification of infected individuals, who may be asymptomatic or oligosymptomatic. Protection of HCWs not only serves as personal protection but is also paramount to mitigate nosocomial transmission to vulnerable patients and fellow HCWs. HCW infections may also lead to critical staff shortages. Various infection prevention and control interventions have been adapted worldwide with various efficacies, including the use of adequate PPE ^6-8^, universal admission screening of patients ^9 10^, and RT-PCR surveillance of exposed HCWs.^11 12^ However, reports about limited availability of sufficient PPE and high infection rates in HCWs, particularly during the early phase of the outbreak, have raised concerns about whether HCWs can be adequately protected from contracting occupational infections.^13 14^ The World Health Organization recently reported that HCWs account for over 10% of overall global covid-19 cases.^15^ While strict adherence to adequate PPE in areas caring for covid-19 patients is of major importance, this alone may be insufficient in protecting HCWs and in preventing nosocomial infections due to the infectious risk posed by as yet unidentified patients and colleagues. The aim of our study was to prospectively evaluate the effectiveness of sequentially instituted multimodal infection control interventions at our large tertiary care centre by sequential seroprevalence measurements and assessment of seroconversion rates amongst the various hospital workers.

## METHODS

### Study design

Participants were recruited in the screening period 1 (SP 1) between 20 March and 9 April 2020 by informing employees of the University Medical Center Hamburg-Eppendorf. The city-state of Hamburg with 1.8 million inhabitants and a metropolitan catchment area of more than 4.5 million inhabitants was affected by the pandemic relatively early due to public holidays in March 2020, when numerous travelers returned from high-risk regions, especially Italian and Austrian ski resorts. By 24 July, a total of 5291 cases had been confirmed, which constitutes one of the highest rates in Germany.^16^ The University Medical Center Hamburg-Eppendorf, which has 1738 hospital beds, has treated more than 170 covid-19 patients during the study period. Of those, 70 were admitted to the intensive care unit, 51 required mechanical ventilation, and 12 patients required extracorporeal membrane oxygenation (ECMO) therapy. Of note, a large number of immunocompromised patients with high viral loads and a protracted course of disease posed a particularly high transmission risk to HCWs.^17^ Participants were recruited to the study via an internal email newsletter. To assess the infection rate of employees, we assessed seroconversion at one and three months: i) sampling period 2 (SP 2) one month after SP 1 (20 April to 8 May 2020) and ii) sampling period 3 (SP 3) another 2 months after SP 2 (22 June to 24 July).

### Data collection

On the day of recruitment, we collected demographic and general work-related data including profession, occupational area, and patient contacts using a questionnaire. In addition, we used another questionnaire to assess known and possible past contact to covid-19 patients with or without sufficient PPE as well as presence of symptoms during the past 4 weeks at SP 1 and SP 2 and during the past 8 weeks at SP 3 respectively. Fever, cough, and dyspnea were classified as typical symptoms while rhinorrhea, sore throat, headache, stomach pain, joint and muscle pain, nausea, and diarrhea and were considered as uncharacteristic symptoms. Questionnaires were available both paper-based and via online REDcap electronic data capture tools hosted at our institution.^18 19^ Study participants with positive SARS-CoV-2 IgG ratio were contacted again by phone to ask if nasopharyngeal swabs for RT-PCR had been performed and whether a source of infection could be identified.

### Infection control interventions

A number of infection control measures were undertaken at our institution listed in table 1.

**Table 1.** Infection control interventions at the University Medical Center Hamburg-Eppendorf and in Hamburg, Germany.

| **Interventions at the University Medical Center Hamburg-Eppendorf** |  |
| --- | --- |
| 27 February | Opening of an on-campus covid-19 testing clinic |
| 11 March | Cancellation of all meetings not directly related to patient care |
| 18 March | Mandatory wearing of face masks in the emergency department |
| 19 March | Implementation of visitor restrictions |
| 22 March | Mandatory wearing of face masks in all clinical settings |
| 10 April | Mandatory wearing of FFP2 masks at all oncology and haematology wards and clinics |
| 20 April | Universal RT-PCR admission screening of patients |
| 11 May | Universal RT-PCR screening of employees caring for covid-19 patients or vulnerable patients |
| **Interventions in Hamburg, Germany** |  |
| 16 March | Closure of educational facilities |
| 22 March | Stay at home order |
| 27 April | Mandatory wearing of face masks |

As a large tertiary referral centre and one of the major infectious diseases units of the country, we have highly trained personnel and a specialized infectious disease ward. In addition, additional covid-19 units were opened both in intensive care and for standard care, following intensive staff training. Thus, patient care with full PPE including FFP2 masks of all identified covid-19 patients was standard of care from the beginning of the pandemic, and even though supply was scarce throughout the first two months, application of strict protocols to limit wasteful use of equipment allowed sufficient supply all through the pandemic. When first infections in staff members occurred, several further steps were undertaken starting with the universal use of face masks in all clinical settings, and measures were intensified further after a nosocomial outbreak on one of the haematology wards.^17^ In addition to separate covid-19 units, use of full PPE in all covid-19 care, and use of surgical face masks in all clinical units of the hospital, universal screening of all hospital admissions was instituted at April 20, with a roll-out period of about ten days until full adherence across all hospital departments. In addition, RT-PCR testing of staff at risk was offered as of May 11, but this measure was primarily instituted as a reassurance measure for our staff rather than an infection control measure.

### Study procedures

During all three sampling periods, serum samples were drawn from the study participants. A semi-quantitative SARS-CoV-2 IgG enzyme-linked immunosorbent assay (ELISA) targeting the S1-Domain of the S-protein spike protein subunit was performed using a commercial kit (Euroimmun Medizinische Labordiagnostika, Lübeck, Germany). Testing procedures were performed according to the manufacturer’s instructions. According to the manufacturer, a ratio of ≥1.1 should be regarded as positive. The manufacturer reports a specificity of 99.3%. We have recently independently validated those results and showed a specificity of 99.1%.^20^ However, an optimized IgG ratio of ≥ 1.5, has been shown to display a specificity of 100% by us and by others.^21^ Therefore, we used both the manufacturer’s and this more stringent cut-off value for positive results to account for optimal specificity in a low prevalence environment. Given the sequential testing in our study population and the expected rise of IgG antibody ratio ≥1.5 at some time point in infected individuals, this cut-off value is considered the best compromise for optimum sensitivity and specificity. Study participants were classified as seropositive in all future sampling periods, if they had an IgG antibody ratio ≥1.5 at least once, even if antibody ratios waned over time. This was found to be the case in three study participants, who became seronegative at SP 3. However, in order to also look at the data with maximum sensitivity, we also analyzed the participants with a ratio of >1.1.

### Statistical analyses

Results were subjected to statistical analysis using SPSS 20.0 (IBM, Armonk, NY, USA). Fishers’ exact test was used to the determine association between two categorical variables. A two-sided P value less than 0.05 was considered significant.

### Patient and public involvement

The study was presented to and approved by the academic and non-academic workers representation boards of our hospital and the design agreed with both boards as well as the management board of the hospital.

## RESULTS

### Study population

A total of 1253 individuals were included during SP 1, which represent around 11% of all employees at our centre (table 2).

**Table 2.** Characterization of the study population.

|  |  |
| --- | --- |
| **n** | 1253 |
| **Sex, n (%)** |  |
| Female | 934 (74.5) |
| Male | 308 (24.6) |
| Diverse | 11 (0.9) |
| **Age** |  |
| Range, years | 16-69 |
| Mean, years | 38.4 |
| **Occupation, n (%)** |  |
| HCW | 1026 (81.9) |
| *Category* |  |
| - Nurse | 444 (35.4) |
| - Physician | 275 (21.9) |
| - Medical technician | 105 (8.4) |
| - Medical student | 73 (5.8) |
| - Physiotherapist | 15 (1.2) |
| - Other | 114 (9.1) |
| *Location* |  |
| - Regular ward | 332 (26.5) |
| - Outpatient clinic | 234 (18.7) |
| - Intensive care unit | 130 (10.4) |
| - Operating room | 111 (8.9) |
| - Emergency department | 63 (5.0) |
| - Other | 156 (12.5) |
| Non-HCW | 227 (18.1) |

All study participants participated at the second follow up visit, but 23 (1.8%) missed the first follow-up visit. The average age of the study cohort was 38.4 years; 934 were women and 308 were men. The majority of participants (n=1026, 81.9%) were HCWs such as nurses (n=444, 35.4%), medical doctors (n=275, 21.9%), medical technicians with regular patient contact (n=105, 8.4%) and physiotherapists (n=15, 1.2%). Altogether, the participants well represented the average workforce of our centre, consisting of 30.1% nurses, 25.8% medical doctors, and 44.1% other staff members, with a somewhat higher proportion of front-line HCWs in the study. Study participants were recruited from all occupational areas of our hospital including regular wards (n=332, 26.5%), outpatient clinics (n=234, 18.7%), the intensive care unit (n=130, 10.4%) and the emergency department (n=63, 5.0%). The remainder of 227 (18.1%) participants not classified as HCWs were research scientists, administrative staff or belonged to other occupational groups not directly involved in patient care. Information on contact with covid-19 was provided by 1253 individuals at SP 1, 1169 individuals at SP 2, and 1110 individuals at SP 3 (table 3).

**Table 3.**
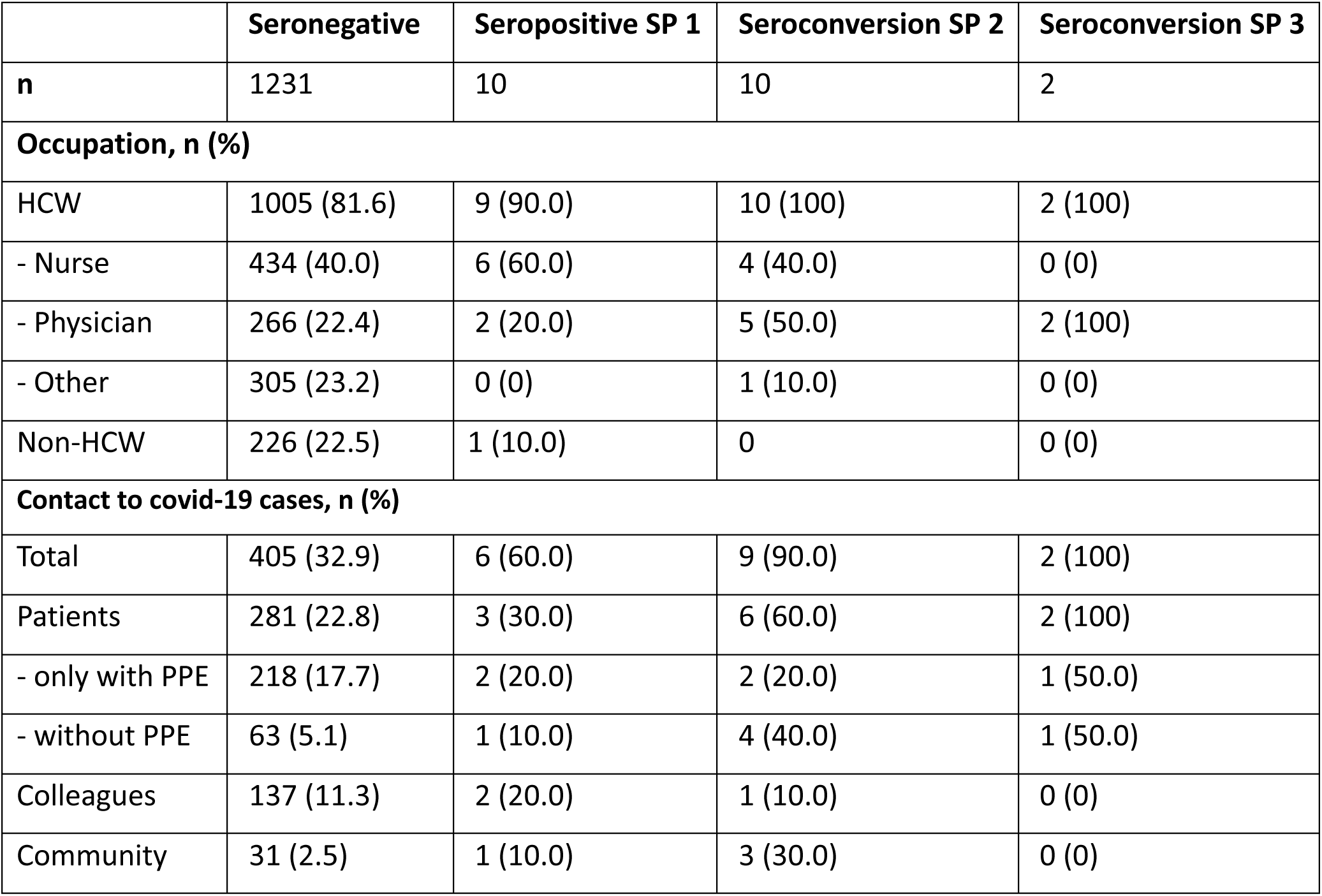
Occupational exposure of the study population.

|  | **Seronegative** | **Seropositive SP 1** | **Seroconversion SP 2** | **Seroconversion SP 3** |
| --- | --- | --- | --- | --- |
| **n** | 1231 | 10 | 10 | 2 |
| **Occupation, n (%)** |  |  |  |  |
| HCW | 1005 (81.6) | 9 (90.0) | 10 (100) | 2 (100) |
| - Nurse | 434 (40.0) | 6 (60.0) | 4 (40.0) | 0 (0) |
| - Physician | 266 (22.4) | 2 (20.0) | 5 (50.0) | 2 (100) |
| - Other | 305 (23.2) | 0 (0) | 1 (10.0) | 0 (0) |
| Non-HCW | 226 (22.5) | 1 (10.0) | 0 | 0 (0) |
| **Contact to covid-19 cases, n (%)** |  |  |  |  |
| Total | 405 (32.9) | 6 (60.0) | 9 (90.0) | 2 (100) |
| Patients | 281 (22.8) | 3 (30.0) | 6 (60.0) | 2 (100) |
| - only with PPE | 218 (17.7) | 2 (20.0) | 2 (20.0) | 1 (50.0) |
| - without PPE | 63 (5.1) | 1 (10.0) | 4 (40.0) | 1 (50.0) |
| Colleagues | 137 (11.3) | 2 (20.0) | 1 (10.0) | 0 (0) |
| Community | 31 (2.5) | 1 (10.0) | 3 (30.0) | 0 (0) |

A total of 23.3% (n=292) of participants reported having been in contact with known covid-19 patients. The majority of those (n=223) worked exclusively in the direct care of patients with diagnosed SARS-CoV-2 infections and thus reported to always be equipped with appropriate PPE. Another 69 HCWs had contact with patients that were diagnosed with covid-19 only later, i.e. after exposure, and did therefore not wear adequate PPE at the time. Occupational contact to infected colleagues was reported by 11.2% (n=140), and known community contacts by 2.8% (n=35) of participants.

### Serological results

At the initial screening period a total of 0.8% (n=10, 95% confidence interval [CI] 0.3 to 1.3) participants were found to be SARS-CoV-2 seropositive (figure 1). Another ten individuals, representing 0.8% of all study participants at SP 2, showed seroconversion at the first follow-up visit, giving a seroprevalence of 1.6% (95% CI 0.9 to 2.3). At SP 3, two more individuals had developed SARS-CoV-2 IgG antibodies, giving a seroconversion rate of 0.16% between SP 2 and SP 3, or 0.08% per month, and thus a total seroprevalence of 1.8% (n=22, 95% CI 1.0 to 2.5) by the end of the study when using the optimized IgG ratio of ≥ 1.5. One of the two participants showing seroconversion only at SP 3 had indeed a positive RT-PCR for SARS-CoV-2 and missed the first follow-up visit since he was in quarantine. While it is likely that that this seroconversion took place before SP 2, we have no proof of this, thus the case was counted as converting in the second observation period.

**Figure 1.**
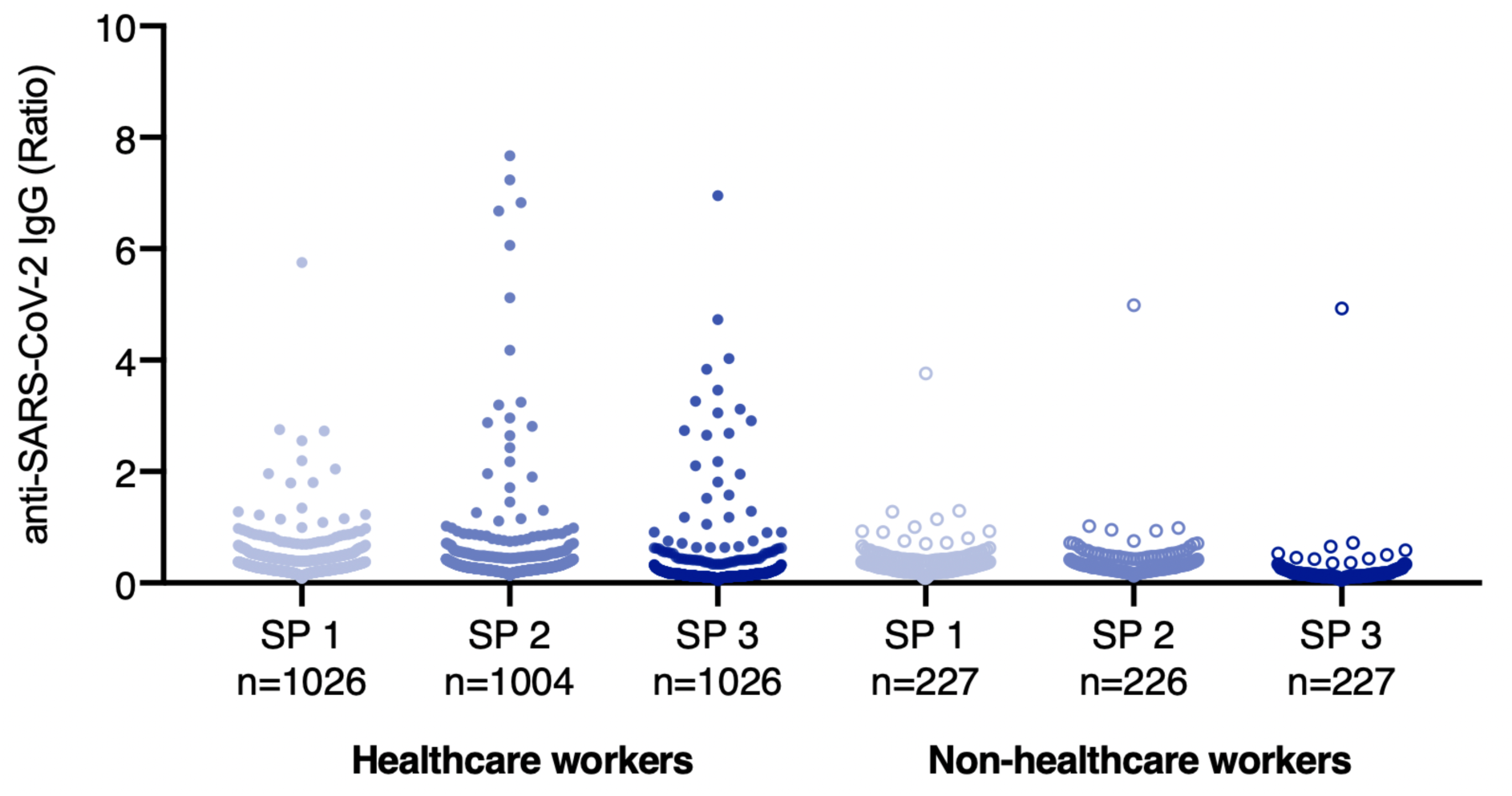
Serologic results of the study population.

Three participants with an IgG ratio of ≥ 1.5 at SP 1 subsequently had a negative test result at SP 2 and SP 3 respectively suggesting that their infection had occurred early. This demonstrates that a cross-sectional study with a single measurement only at SP 3 would have missed these three infections. When analyzing the serological data with of maximum sensitivity, i.e. with a cut-off value of ≥ 1.1, the overall results were very similar: 19 participants showed anti-SARS-CoV2 antibodies with a ratio ≥ 1.1 at SP 1, another 13 participants showed seroconversion during the first month between SP 1 and SP 2. Only 2 additional participants showed seroconversion with this lower cut-off in the subsequent two months, bringing the total of possible seroconverters in the last two months up to four, or two per month (figure 2).

**Figure 2.**
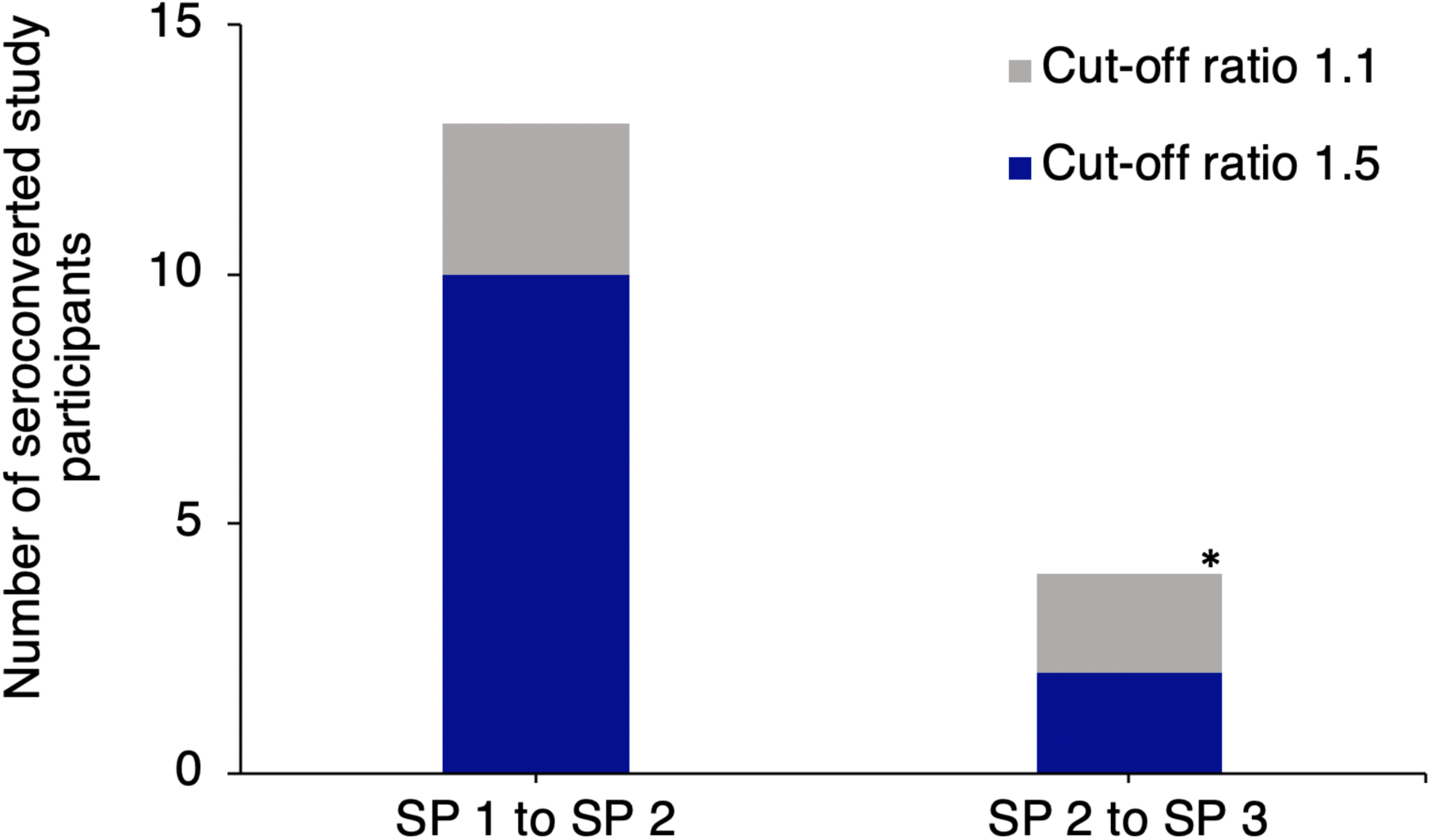
Seroconversion rates in study participants using different cut-off ratios. *One of the study participants with seroconversion between SP 2 and SP 3 had been tested positive for SARS-CoV-2 by RT-PCR between SP 1 and SP 2, did therefore not attend the first follow-up visit, and was thus counted as seroconverting between SP 2 and SP 3.

### RT-PCR testing history of seroconverted participants

Out of all clearly seropositive study participants, a total of 40.9% (n=9), one of them before SP 1 and the others between SP 1 and SP 2, had been diagnosed with covid-19 by RT-PCR and thus had been placed under quarantine during the study period. Another six seropositive HCWs reported that they had at least one nasopharyngeal swab performed due to symptoms or close contact with a covid-19 case but were tested negative at that time. The remainder of seven clearly seropositive individuals had not been tested by RT-PCR at all.

### Risk factors for seroconversion

Complete information on symptoms throughout the study period was provided by 1061 (84.7%) of the study participants. Of those, 54.9% (n=582) reported covid-19 related symptoms (see supplementary table). Only 10.6% (n=112) did not report any symptoms throughout the study period, the rest reported symptoms not considered typical for covid-19. Typical SARS-CoV-2 symptoms were reported by 72.7% (n=16) of seropositive and 54.4% (n=566) of seronegative study participants (P=0.4). The probable source of infection could be identified for the majority of seropositive study participants. A total of six HCWs presumably got infected when caring for patients that were only subsequently diagnosed with covid-19 and had thus not used appropriate PPE at the time of exposure. Five HCWs most likely contracted SARS-CoV-2 while caring for covid-19 patients with adequate PPE. Other probable infection routes in our cohort were close contact with infected colleagues (n=3), community transmission in high-risk regions (n=3), and infected household contacts (n=1). For the remainder of four seropositive employees, the source of infection could not be established. Seropositivity was not statistically significantly higher in HCWs (2.0%, n=21/1026) compared to other hospital workers (0.4%, n=1/227, P=0.16). When only addressing HCWs directly involved in the care of patients with SARS-CoV-2 infections (3.8%, n=11/292) compared to other HCWs (1.4%, n=10/734, P=0.025) seropositivity was significantly increased. Importantly, both seroconverters of SP3 were not involved in inpatient care, but exclusively worked in the outpatient clinics.

## DISCUSSION

This study demonstrates a very low overall SARS-CoV-2 seroprevalence in hospital workers at our tertiary care centre, and the increasing effectiveness of a combination of infection control measures. The low overall in-hospital SARS-CoV-2 transmission rates of healthcare personnel were achieved despite both substantial community transmission in the city-state of Hamburg, and, more importantly, a large number of covid-19 patients treated, some with very high and long-lasting viral loads, at our tertiary care centre. While the relatively early detection of the first cases in Germany ^22^ may have increased preparedness, the addition of further multimodal infection prevention and control measures was instrumental in minimizing occupational exposure risks. While seropositivity amongst HCWs was initially higher in HCWs directly involved in the care of patients with SARS-CoV-2 infections compared to other employees, only two more seroconversions were observed after the implementation of all multimodal infection during the last two months, and both of these did not involve inpatient care but seemed to have occurred during outpatient clinics or from the community. Also, one of those two physicians had been tested positive for SARS-CoV-2 by RT-PCR between SP 1 and SP 2 and did therefore not attend the first follow-up visit, so that this seroconversion may also have also occurred the first observation period. Thus, presumably only one study participant, a physician employed in an outpatient clinic, contracted a SARS-CoV-2 infection in the two months between SP 2 and SP 3. All other participants were protected despite a considerable number of covid-19 patients being treated at our centre during that time period (figure 3). Considerably higher seroprevalence has been reported for HCWs in Belgium (6,4 – 12,6%) ^23 24^, Spain (11 – 31,6%) ^25 26^, the United Kingdom (24 - 43.5%) ^27 28^ and the United States of America (7,6 - 36%) ^29 30^. Those high infection rates are likely to be at least partially attributable to insufficient protection of exposed HCWs. While the effectiveness of universal use of face masks at hospitals has been shown to significantly decrease hospital transmission ^6 7^, shortages of appropriate PPE for HCWs in many hospitals have emerged particularly during the early phase of the pandemic. However, adequate PPE when caring for confirmed covid-19 patients alone is not sufficient to prevent healthcare transmission, since asymptomatic and pre-symptomatic individuals can play a pivotal role in the transmission of SARS-CoV-2.^4^ Also at our centre, the repeated detection of SARS-CoV-2 positive yet asymptomatic patients presented an important exposure risk leading to occupational infections. Systematic RT-PCT testing of all hospital admissions, either prior to scheduled admissions or in the emergency department, instituted at the end of April, reduced this infection risk to practically zero. The fact that three HCWs seroconverted during the study period despite reporting use of appropriate PPE when caring for covid-19 patients underlines that PPE alone is not sufficient to prevent viral transmission to HCWs. To our surprise, the majority of seropositive study participants had not been diagnosed with SARS-CoV-2 infections despite a very low threshold in our hospital for RT-PCR screening, with more than 7.300 nasopharyngeal swab examinations of hospital HCWs performed during the study period. Furthermore, symptoms compatible with covid-19 were widely prevalent in healthcare professionals regardless of serological results, a number of seropositive employees did not report any such symptoms and were thus not tested. This demonstrates that symptom-based testing of HCWs, while an important tool to mitigate nosocomial infections, has limited sensitivity and very low specificity in detecting infected HCWs. Complete protection of HCWs is not only helpful for their direct protection but is also key for preventing nosocomial infections in patients.

**Figure 3.**
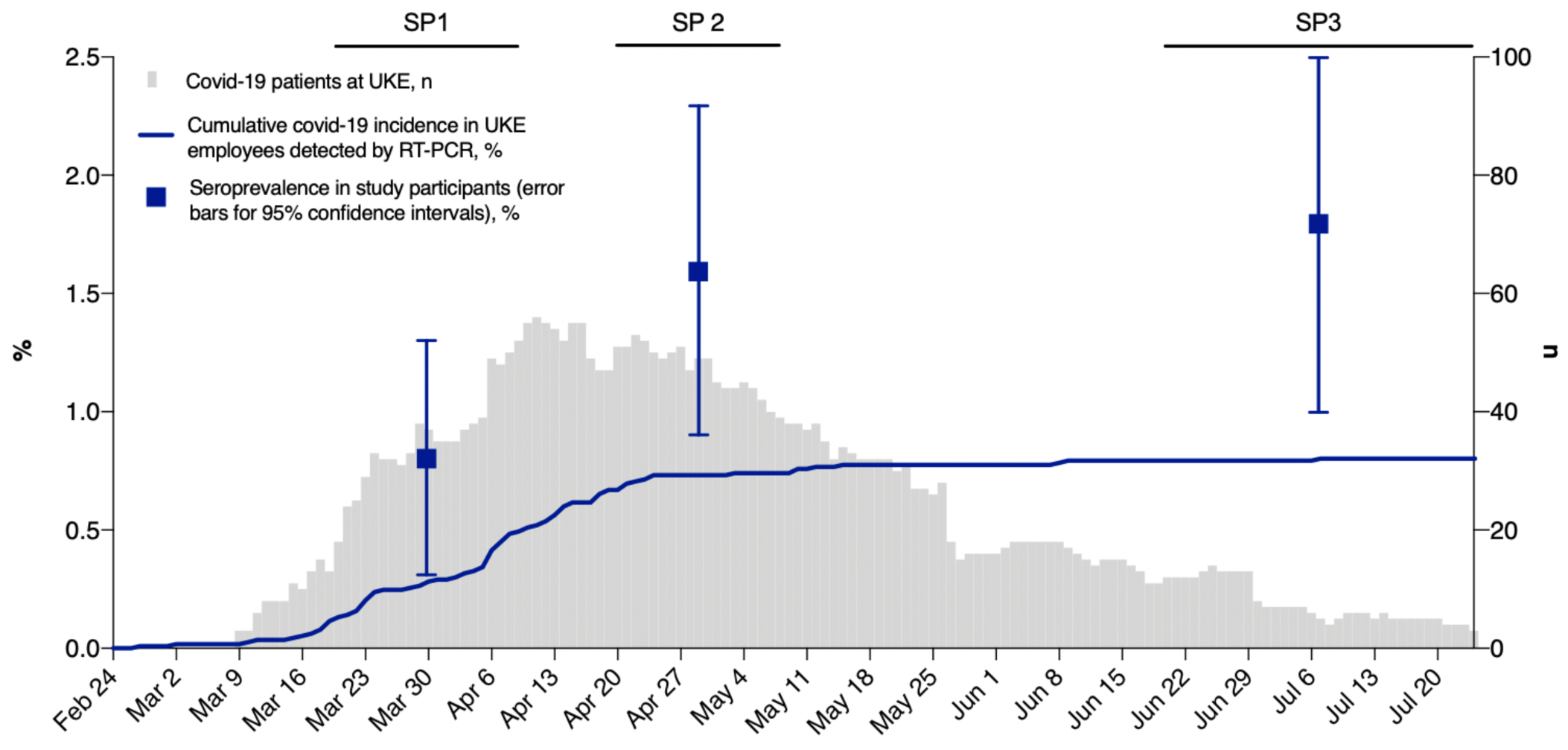
Timeline of the covid-19 epidemic at the University Medical Center Hamburg-Eppendorf and in Hamburg, Germany. Grey bars represent the number of covid-19 patients hospitalized at the University Medical Center Hamburg-Eppendorf (UKE) on the respective days. The blue line represents the cumulative covid-19 incidence in the 11.348 employees of the UKE detected by the low threshold contact- and symptom-based RT-PCR screening. The seroprevalence in the study cohort at the respective sampling periods is represented by blue squares with error bars for 95% confidence intervals.

## Limitations

There are some limitations to our study: firstly, we did not recruit a strictly representative sample of healthcare professionals at our institution, since participation was voluntary. However, the distribution of professions was close to the overall hospital workforce distribution. We included a somewhat higher proportion of clinically exposed HCWs, in particular front-line HCWs from the emergency department, intensive care and infectious diseases departments, most likely because these HCWs were highly exposed and thus more motivated to participate in the study. Secondly, since several multimodal infection control interventions were sequentially adopted and the study did not include a control group, this study is not able to answer the question which measures are most effective in preventing healthcare transmission, and which might have been superfluous. Thirdly, sensitivity and specificity of serological assays is limited, so a precise assessment of the number of infected HCWs is not possible. However, the results were independent of the cut-off levels used for serological testing (figure 2), supporting the reliability of the results.

## Conclusion

We report a very low seroprevalence amongst HCWs and other healthcare professionals at our tertiary care centre at the beginning of the covid-19 epidemic and very few seroconversions after the implementation of multimodal infection prevention and control interventions. When in addition to personal protection and universal face masks RT-PCR screening of hospital admissions was instituted, occupational transmission could be brought down to zero cases in inpatient care and only one or two related to outpatient clinics. Taken together, our findings demonstrate that multimodal infection control measures are able to effectively prevent healthcare transmission for SARS-CoV-2, and thus protect both HCWs and patients.

## Data Availability

No additional data available.

## Contributors

TTB, DS, JSzW, and AWL contributed equally to this paper. TTB, DS, JSzW, and AWL conceived and designed the study. TTB, SL, VS, JK, MT, and FU contributed to recruitment of healthcare professionals, data collection and data analysis. DS, ML and JKK contributed to literature search, data collection and data analysis. TTB and DS contributed to data visualization. SH, SS, MMA, ML, and JKK contributed to data interpretation. TTB, JSzW, and AWL drafted the manuscript. DS, SH, SS and ML reviewed and edited the manuscript. TTB, DS, JSzW, and AWL are the guarantors. The corresponding author attests that all listed authors meet authorship criteria and that no others meeting the criteria have been omitted

## Transparency declaration

The guarantors affirm that this manuscript is an honest, accurate, and transparent account of the study being reported; that no important aspects of the study have been omitted; and that any discrepancies from the study as planned (and, if relevant, registered) have been explained.

## Ethical approval

The study protocol was reviewed and approved by the Ethics Committee of the Medical Council of Hamburg (PV 7298). Written informed consent was obtained by all study participants prior to enrolment.

## Dissemination to participants and related patient and public communities

The findings of this study will be disseminated to all departments of the University Medical Center Hamburg-Eppendorf and to all employees of our institution via an internal newsletter.

## Acknowledgments

We thank all study participants and departments of the University Medical Centre Hamburg-Eppendorf for active participation in the study. We thank Sabrina Kreß, Jennifer Wigger, Martina Schulz, Corinna Eggers, Angelika Schmidt, Silke Kummer, Robin Woost, Nils Dittberner, and Marcus Wurlitzer for excellent technical assistance.

## Data sharing

No additional data available.

---

**Supplementary table.** Symptoms reported by seropositive and -negative study participants. Respective symptoms as stated by the study participants at SP 1 or SP 2 for the preceding month and at SP 3 for the preceding 2 months. Complete information was provided by a total of 1061 participants.

|  | Total (n=1061) | Seropositive (n=22) | Seronegative (n=1041) |
| --- | --- | --- | --- |
| Any symptoms, n(%) | 949 (89.4) | 20 (90.9) | 929 (89.1) |
| Typical symptoms, n(%) | 582 (54.9) | 16 (72.7) | 566 (54.4) |
| Fever, n(%) | 112 (10.6) | 8 (36.4) | 104 (1.0) |
| Cough, n(%) | 498 (46.9) | 11 (50.0) | 487 (46.8) |
| Dyspnea, n(%) | 231 (21.8) | 9 (40.9) | 222 (21.3) |
| Rhinorrhea, n(%) | 717 (67.7) | 17 (77.3) | 700 (67.2) |
| Sore throat, n(%) | 560 (52.8) | 11 (50.0) | 549 (52.7) |
| Headache, n(%) | 704 (66.4) | 16 (72.7) | 688 (66.1) |
| Abdominal pain, n(%) | 295 (27.8) | 8 (36.4) | 287 (27.6) |
| Muscle aches, n(%) | 244 (23.0) | 13 (59.1) | 231 (22.2) |
| Nausea, n(%) | 213 (20.1) | 8 (36.4) | 205 (19.7) |
| Diarrhea, n(%) | 296 (27.9) | 8 (36.4) | 288 (27.7) |

## References

1. Zhan M, Qin Y, Xue X, et al. Death from Covid-19 of 23 Health Care Workers in China. N Engl J Med 2020 doi: 10.1056/NEJMc2005696

2. Remuzzi A, Remuzzi G. COVID-19 and Italy: what next? Lancet 2020;395(10231):1225–28. doi: 10.1016/s0140-6736(20)30627-9

3. Petersen E, Hui D, Hamer DH, et al. Li Wenliang, a face to the frontline healthcare worker. The first doctor to notify the emergence of the SARS-CoV-2, (COVID-19), outbreak. Int J Infect Dis 2020;93:205–07. doi: 10.1016/j.ijid.2020.02.052

4. Arons MM, Hatfield KM, Reddy SC, et al. Presymptomatic SARS-CoV-2 Infections and Transmission in a Skilled Nursing Facility. N Engl J Med 2020;382(22):2081–2090. doi: 10.1056/NEJMoa2008457

5. Gandhi M, Yokoe DS, Havlir DV. Asymptomatic Transmission, the Achilles’ Heel of Current Strategies to Control Covid-19. N Engl J Med 2020 doi: 10.1056/NEJMe2009758

6. Liu M, Cheng SZ, Xu KW, et al. Use of personal protective equipment against coronavirus disease 2019 by healthcare professionals in Wuhan, China: cross sectional study. BMJ 2020;369:m2195. doi: 10.1136/bmj.m2195

7. Wang X, Ferro EG, Zhou G, et al. Association Between Universal Masking in a Health Care System and SARS-CoV-2 Positivity Among Health Care Workers. JAMA 2020 doi: 10.1001/jama.2020.12897

8. Pan A, Liu L, Wang C, et al. Association of Public Health Interventions With the Epidemiology of the COVID-19 Outbreak in Wuhan, China. JAMA 2020;323(19):1–9. doi: 10.1001/jama.2020.6130

9. Sutton D, Fuchs K, D’Alton M, et al. Universal Screening for SARS-CoV-2 in Women Admitted for Delivery. N Engl J Med 2020;382(22):2163–64. doi: 10.1056/NEJMc2009316

10. Al-Shamsi HO, Coomes EA, Alrawi S. Screening for COVID-19 in Asymptomatic Patients With Cancer in a Hospital in the United Arab Emirates. JAMA Oncol 2020 doi: 10.1001/jamaoncol.2020.2548

11. Treibel TA, Manisty C, Burton M, et al. COVID-19: PCR screening of asymptomatic health-care workers at London hospital. The Lancet 2020 doi: 10.1016/s0140-6736(20)31100-4

12. Black JRM, Bailey C, Przewrocka J, et al. COVID-19: the case for health-care worker screening to prevent hospital transmission. Lancet 2020;395(10234):1418–20. doi: 10.1016/s0140-6736(20)30917-x

13. Ranney ML, Griffeth V, Jha AK. Critical Supply Shortages - The Need for Ventilators and Personal Protective Equipment during the Covid-19 Pandemic. N Engl J Med 2020;382(18):e41. doi: 10.1056/NEJMp2006141

14. Wang J, Zhou M, Liu F. Reasons for healthcare workers becoming infected with novel coronavirus disease 2019 (COVID-19) in China. J Hosp Infect 2020;105(1):100–01. doi: 10.1016/j.jhin.2020.03.002

15. World Health Organization (WHO) Coronavirus disease (COVID-2019) press briefings. https://www.who.int/docs/default-source/coronaviruse/transcripts/covid-19-virtual-press-conference17-july.pdf?sfvrsn=dd7f91a1_0 [Accessed on 29 July 2020]

16. Robert Koch Institute. Coronavirus Disease 2019 (COVID-19) Daily Situation Report of the Robert Koch Institute. https://www.rki.de/DE/Content/InfAZ/N/Neuartiges_Coronavirus/Situationsberichte/2020-07-27-en.pdf?__blob=publicationFile [Accessed on 29 July 2020]

17. Weisel KC, Morgner-Miehlke A, Petersen C, et al. Implications of SARS-CoV-2 Infection and COVID-19 Crisis on Clinical Cancer Care: Report of the University Cancer Center Hamburg. Oncol Res Treat 2020;43(6):307–13. doi: 10.1159/000508272

18. Harris PA, Taylor R, Thielke R, et al. Research electronic data capture (REDCap)--a metadata-driven methodology and workflow process for providing translational research informatics support. J Biomed Inform 2009;42(2):377–81. doi: 10.1016/j.jbi.2008.08.010

19. Harris PA, Taylor R, Minor BL, et al. The REDCap consortium: Building an international community of software platform partners. J Biomed Inform 2019;95:103208. doi: 10.1016/j.jbi.2019.103208

20. Pflüger LS, Bannasch JH, Brehm TT, et al. Clinical evaluation of five different automated SARS-CoV-2 serology assays in a cohort of hospitalized COVID-19 patients. J Clin Virol 2020:104549. doi: 10.1016/j.jcv.2020.104549

21. Meyer B, Torriani G, Yerly S, et al. Validation of a commercially available SARS-CoV-2 serological Immunoassay. medRxiv 2020:2020.05.02.20080879. doi: 10.1101/2020.05.02.20080879

22. Stafford N. Covid-19: Why Germany’s case fatality rate seems so low. BMJ 2020;369:m1395. doi: 10.1136/bmj.m1395

23. Steensels D, Oris E, Coninx L, et al. Hospital-Wide SARS-CoV-2 Antibody Screening in 3056 Staff in a Tertiary Center in Belgium. JAMA 2020 doi: 10.1001/jama.2020.11160

24. Martin C, Montesinos I, Dauby N, et al. Dynamic of SARS-CoV-2 RT-PCR positivity and seroprevalence among high-risk health care workers and hospital staff. J Hosp Infect 2020 doi: 10.1016/j.jhin.2020.06.028

25. Garcia-Basteiro AL, Moncunill G, Tortajada M, et al. Seroprevalence of antibodies against SARS-CoV-2 among health care workers in a large Spanish reference hospital. medRxiv 2020:2020.04.27.20082289. doi: 10.1101/2020.04.27.20082289

26. Galan I, Velasco M, Casas ML, et al. SARS-CoV-2 seroprevalence among all workers in a teaching hospital in spain: unmasking the risk. medRxiv 2020:2020.05.29.20116731. doi: 10.1101/2020.05.29.20116731

27. Houlihan C, Vora N, Byrne T, et al. SARS-CoV-2 virus and antibodies in front-line Health Care Workers in an acute hospital in London: preliminary results from a longitudinal study. medRxiv 2020:2020.06.08.20120584. doi: 10.1101/2020.06.08.20120584

28. Shields AM, Faustini SE, Perez-Toledo M, et al. SARS-CoV-2 seroconversion in health care workers. medRxiv 2020:2020.05.18.20105197. doi: 10.1101/2020.05.18.20105197

29. Mansour M, Leven E, Muellers K, et al. Prevalence of SARS-CoV-2 Antibodies Among Healthcare Workers at a Tertiary Academic Hospital in New York City. J Gen Intern Med 2020:1–2. doi: 10.1007/s11606-020-05926-8

30. Stubblefield WB, Talbot HK, Feldstein L, et al. Seroprevalence of SARS-CoV-2 Among Frontline Healthcare Personnel During the First Month of Caring for COVID-19 Patients - Nashville, Tennessee. Clin Infect Dis 2020 doi: 10.1093/cid/ciaa936

